# Interventions for improving antibiotic prescribing in the community: systematic review and meta-analysis

**DOI:** 10.1101/2024.11.20.24317684

**Authors:** Rebecca A J Andrews, Virginia Hernandez Santiago, Bruce Guthrie, Charis A Marwick

## Abstract

**Background:** Antibiotic resistance is a public health priority and antibiotic use in humans is a major contributing factor to its development. Interventions to improve antibiotic prescribing in the community, where most antibiotics are prescribed, are widely implemented with varying effect. The aim was to systematically review and meta-analyse evidence on effectiveness of different types of antibiotic prescribing interventions in the community.

**Methods and Findings:** Medline, Embase, and the Cochrane Central Register of Controlled Trials, were searched from database inception to 16 August 2021 to identify randomised controlled trials comparing antibiotic stewardship interventions versus usual care in community settings. Two reviewers screened studies, extracted data, and assessed risk of bias. Studies were grouped according to type of intervention. Meta-analyses employed random effects models. The outcome for meta-analyses was change in total antibiotic prescribing rates attributable to the intervention, compared to usual care, calculated as percentage differences. Other measures of change in antibiotic prescribing were included in narrative description.

Of 26,130 studies screened, 74 were included, with 53 comparisons from 45 studies meta- analysed. 50% of included studies had high risk of bias. Single interventions with statistically significant reductions in total antibiotic prescribing were point of care tests for antigen detection (−28.0% reduction, 95%CI−38.2 to−17.8); educational materials (−17.0%, −31.0 to - 3.0); printed decision-support systems (−10.8%,−15.7 to -6.0), educational workshops (− 8.7%,−12.8 to -4.7), and; educational outreach (−6.0%,−10.6 to−1.4). Multifaceted interventions were not more effective than single interventions (education + audit and feedback –9.9%, −12.8 to -7.0; other multifaceted -9.4%, −17.2 to −1.6). Effect sizes in sensitivity analyses excluding trials at high risk of bias were similar or larger.

**Conclusions:** Community antibiotic stewardship interventions were effective but with considerable variation in effect size. The most effective trial interventions may be more difficult to implement in practice, a key challenge for antibiotic stewardship.

**Systematic review registration** PROSPERO CRD42014010160

## Introduction

Antimicrobial resistance (AMR) is an increasing global public health threat,^1^ which causes significant morbidity, mortality, and healthcare costs.^2, 3^ Predictions published in 2016, of 10 million annual deaths by 2050,^4^ are consistent with statistical models using real-world data which estimated 4·95 million (95% CI 3·62–6·57) deaths associated with bacterial AMR in 2019, with 1·27 million (95% CI 0·91–1·71) attributable to AMR.^5^ Drivers of AMR are multifactorial and include antimicrobial use in animals and agriculture, but inappropriate use and overuse of antibiotics in humans is a major contributing factor. Stopping antibiotic use results in the exponential decay of bacterial resistance at the individual microbiome level,^6^ and impacts on resistance rates in serious bacterial infection at the human population level. ^7^ Reasons behind inappropriate use or overuse of antibiotics are complex, where practitioner, patient, societal, and healthcare system factors all interplay.^8, 9^ Antimicrobial stewardship is the coordinated set of actions designed to promote appropriate use of antimicrobials.^10^ National and global actions to increase stewardship include the UK AMR Global Action Plan 2019-2024 focusing on reducing unnecessary exposure to antibiotics and WHO GLASS system providing a world-wide platform for surveillance of antibiotic use.^11, 12^

Community practice or “primary care” is where most antibiotics are prescribed,^13^ with up to 50% of prescriptions considered inappropriate.^14–16^ Diagnostic uncertainty and fear of complications contributes,^17^ along with lack of clinician awareness of the contribution of primary care antibiotic use to AMR.^9^ Previous literature ^18^ including systematic reviews of clinician-targeted interventions for improving antibiotic use in inpatient^19^ and outpatient settings^16^ found that interventions using professional education, feedback, clinical decision support and delayed prescribing have some effect, and multi-component interventions may be more effective.^16^ However, the most recent systematic review in the community including a comprehensive range of intervention and infection types is over 15 years old, and the included studies were too heterogenous to meta-analyse, ^16^ limiting quantitative assessment and comparison of effectiveness. Narrower systematic reviews report that educational interventions, mainly implementing guidelines, can be effective in general^20^ and for respiratory tract infections,^21^ and that point of care testing using C-reactive protein (CRP) for respiratory tract infections may also be effective.^22, 23^ Stewardship interventions should, ideally, encompass the broader clinical context and results from single intervention or infection type cannot necessarily be extrapolated to other indications. There is insufficient up-to-date evidence on which components make a primary care stewardship intervention more effective.

The aim of this study is to systematically review and meta-analyse randomised controlled trials of interventions to improve primary care antibiotic prescribing to estimate their effectiveness and examine which intervention types are more effective.

## Methods

The review was registered on PROSPERO^24^ and is reported according to the preferred reporting items for systematic reviews and meta-analyses (PRISMA) statement^25^.

### Eligibility criteria

Studies were eligible if they were randomised clinical trials or cluster randomised trials involving practitioners who prescribe antibiotics in community settings. Community settings included family and general practice, community paediatrics, nursing and residential homes, and clinics attached to hospitals if they were described as a primary care clinic (which is the case in the US). Studies which only involved other hospital clinics or emergency departments were excluded. Interventions which were only patient focussed or public mass media campaigns were not eligible.

### Data sources and search strategy

MEDLINE and EMBASE databases were searched to identify all randomised controlled clinical trials evaluating interventions for improving antibiotic prescribing published on or before 16^th^ August 2021. The Cochrane EPOC Specialised Register was searched for the terms antibiotic* or antimicrobial* in all fields. There were no language restrictions. The complete search strategy is in Supplementary Table S1. The search did not specify healthcare setting, with community interventions identified during screening. Additional studies were identified from bibliographies of included articles.

### Outcome measure

Included studies could measure antibiotic prescribing in various ways, including the decision to prescribe an antibiotic, the choice of which antibiotic to prescribe, the dose or duration of a prescription, measured as changes in total or targeted prescribing, ‘appropriate’ prescribing, compliance with guidelines, or cost.

For meta-analyses, the outcome measure was the difference in total antibiotic prescribing rates in the intervention group, compared to usual care, calculated as the mean percentage difference. This was either the mean difference between intervention and control groups at follow-up or, using the mean change score between baseline and follow-up for each group, the difference in mean change scores between the groups. The latter was preferred if it was reported or could be calculated. Other measures were described in narrative synthesis.

### Study selection and data extraction

Titles and abstracts were independently screened by two reviewers. Full text articles were appraised by two of four reviewers (RA, VS, BG, CM), after a calibration exercise agreeing common definitions, and inclusion and exclusion rules. Disagreements or uncertainties were resolved by a third reviewer. Studies were grouped by similarity of intervention types based on a modified version of the Cochrane Effective Practice and Organisation of Care (EPOC) Group taxonomy of healthcare interventions,^26^ which included educational interventions, audit and feedback, reminders including decision support systems (DSS), and “other”. For meta-analysis purposes, studies were further categorised into: educational materials, educational workshops, and educational outreach; audit and feedback; computerised DSS, and printed DSS; point of care testing (POCT) using antigen detection (throat swabs for streptococci or influenza), and POCT using inflammatory markers (finger-prick blood tests for C-reactive protein (CRP) or procalcitonin), and; multifaceted interventions, sub- categorised into education plus audit and feedback, and “other” (where there was no apparent dominant intervention). Studies which could not be meta-analysed and/or did not fit into any group were included in narrative description.

Data extraction was performed by one of two reviewers (RA, VHS) and checked by a second reviewer if required. Factorial trials or multiple active arms of a trial were treated as independent comparisons (*versus* standard care) provided there was no statistically significant interaction reported.

### Synthesis of results

Studies were included in meta-analysis if they reported the difference in total antibiotic prescribing rates between arms in a format that could be synthesised (or provided data which allowed calculation into that format). The measure for meta-analysis was the mean percentage difference in total antibiotic prescribing between intervention and control groups at follow-up, or the difference in the mean change from baseline to follow-up between groups. The latter was preferred if it was reported or could be calculated. Meta-analyses were conducted in RevMan 5.4.1 using a random-effects model, which implements a Mantel-Haenszel method and a generic inverse variance method.^27^ Sensitivity analyses excluded studies with high risk of bias.

### Risk of bias

Methodological quality and risk of bias were evaluated using the 2013 Cochrane Effective Practice and Organisation of Care (EPOC) tool (Supplementary Table S2),^26^ by two reviewers (VHS, RA) after a calibration exercise (with BG, CM, who also reviewed cases of discrepancy or doubt). Overall risk of bias was scored ‘Low’ if all criteria were scored as low risk of bias, ’Medium’ if one or two criteria were scored as unclear or high, and ’High’ if more than two criteria were scored as unclear or high.

### Registration

This review was registered in PROSPERO, international prospective register of systematic reviews, registration number CRD42014010160. Deviations from the original protocol with justification are reported in Supplementary Table S3.

## Results

### Included studies

Searches returned 26,130 studies of which 25,748 were excluded during title and abstract screening, with 382 full text articles assessed. 74 studies were included, with 53 comparisons from 45 studies included in meta-analyses (Figure 1, Table 1).

**Figure 1.**
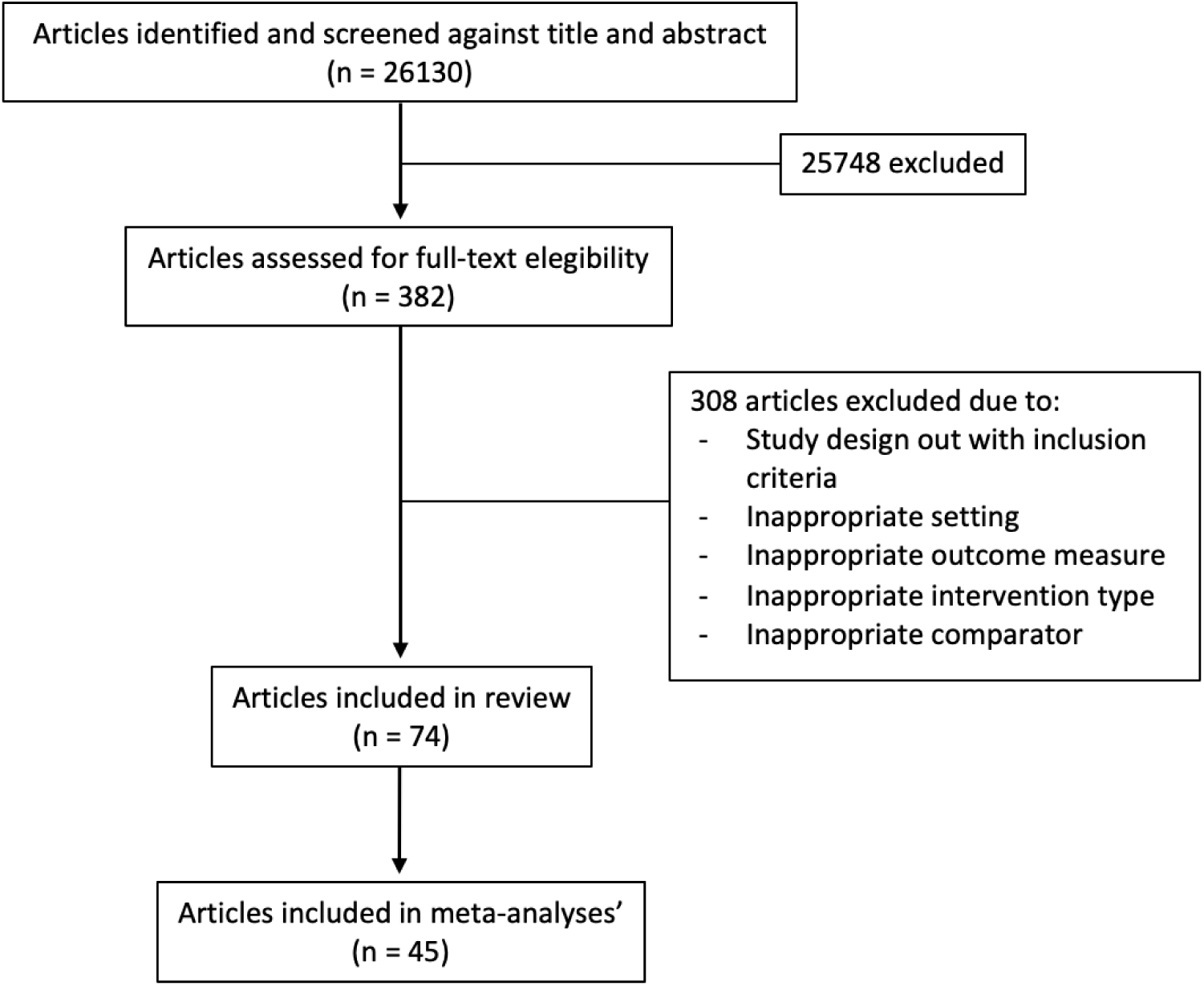
- PRISMA of Included Studies

**Table 1.** Characteristics of Included Studies.

| Study | Country | Population | Intervention | Comparator | Outcome extracted | Overall Risk of Bias | Intervention Group | Meta-analysed |
| --- | --- | --- | --- | --- | --- | --- | --- | --- |
| Altiner (2007) | Germany | Patients with acute cough | Communication skills training through education by peers | Usual care | Total antibiotic use, measured as the rate of antibiotic prescriptions for acute cough by GP | Low | Educational Outreach | No |
| Andreeva (2014) | Russia | Adults with lower respiratory tract infection | Point of Care Testing C-Reactive Protein | Usual care | Rate of total antibiotic prescribing measured as percentage of patients prescribed | High | POCT Inflammatory Markers | Yes |
| Angunawela (1991) | Sri Lanka | All patients | 3 arm trial (2 active arms):<br>- Printed educational material<br>- Face to face education | Usual care | Total antibiotic use, measured as the rate (%) of patients prescribed antibiotics | High | Educational Material<br>Educational Workshop | Yes |
| Arnold (2021) | Denmark | Nursing home residents aged 65 and over | Educational workshop consisting of interactive educational session for nursing staff, use of a dialogue tool containing a checklist for symptoms and a flow chat, and use of a communication tool | Usual care | Antibiotic prescriptions for UTI presented as rate ratios | High | Educational Workshop | No |
| Arroll (2002) | New Zealand | All patients | Delayed prescribing | Usual care | Percentage of patients using antibiotics | Unclear | Other | No |
| Avorn (1983) | U.S. | All patients | 3 arm trial (2 active arms):<br>- Printed educational material<br>- Face to face education<br>- Usual care | Usual care | Specific antibiotic use (cephalexin) measured as mean number of units prescribed per physician | Unclear | Educational Material<br>Educational Outreach | No |
| Awad (2006) | Sudan | All patients | Four arm trial (3 active arms):<br>- Audit and feedback<br>- Audit and feedback plus educational workshop<br>- Audit and feedback plus educational outreach | Usual care | Total antibiotic prescribing measured as mean number of encounters with antibiotic prescribed | Unclear | Audit and Feedback<br>Multifaceted – Education with Audit and Feedback | Yes |
| Bexell (1996) | Sweden | All patients | 3 education seminars within a period of 4 months | Usual care | Total antibiotic use, measured as the rate (%) of patients prescribed antibiotics | Unclear | Educational Workshop | Yes |
| Bourgeois (2010) | U.S. | Children younger than 18 years old with acute respiratory tract infection | Interactive template for acute respiratory tract infections (called 'ARI-IT') | Usual care | Total antibiotic use measured as the percentage of visits with antibiotic prescribed | Unclear | Computerised DSS | Yes |
| Briel (2006) | Switzerland | Patients aged 18 years old or older with acute respiratory tract infection | Education about guidelines | Usual care | Total antibiotic use, measured as the dispensing of antibiotic prescriptions within 2 weeks of consultation, as reported by pharmacists | High | Educational Workshop | Yes |
| Briel (2008) | Switzerland | Patients of any age with acute respiratory tract infection | Point of Care Testing - Procalcitonin | Usual care | Rate of total antibiotic prescribing measured as percentage of patients prescribed | Unclear | POCT Inflammatory Markers | Yes |
| Burkhardt (2010) | Germany | Adults with acute respiratory tract infection | Point of Care Testing- Procalcitonin | Usual care | Total antibiotic prescribing, measured as rate of patients receiving antibiotics | Unclear | POCT Inflammatory Markers | Yes |
| Butler (2012) | UK | All patients | STAR educational programme:<br>- Seminar about own prescribing and resistance<br>- Online education<br>- Consultation skills | Usual care | Total antibiotic use, measured as items/1000 registered patients | Low | Educational Workshop | Yes |
| Cals (2009) |  | Patients with low respiratory tract infection | 2x2 factorial trial:<br>- POCT - CRP<br>- Educational<br>- POCT - CRP with education<br>- Usual care | Usual care | Total antibiotic use, measured as the prescribing rate at index consultation and 1-28 days | Unclear | Educational Workshop<br>POCT Inflammatory Markers | Yes |
| Cals (2013) | The Netherlands | Follow up of an included study (see Cals 2009) | Three interventions were carried out: intervention A included physician use of CRP testing; intervention B included physician communication skills training; and intervention C included both intervention A and B combined | Usual care | The main outcome measure was the average number of episodes of respiratory tract infections during follow up for which patients visited their family physicians for per year<br>The secondary outcome was the percentage of episodes for which patients were treated with antibiotics during follow up | Unclear | Other | No |
| Chazan (2007) | Israel | All patients | One monthly educational seminar over two years versus sessional education | Seasonal education | Total antibiotic use expressed as DDDs/1000 patients/day | Unclear | Other | No |
| Chen (2014) | China | Patients with respiratory tract infections | Use of text messages for guideline dissemination | Usual care | Total antibiotic use measured as number of antibiotic prescriptions (% total) | High | Other | No |
| Christakis (2001) | U.S. | Paediatric population with acute otitis media (AOM) | Evidence presented to providers at point of care through screens | Usual care | Total antibiotic use measured as percentage difference in number of antibiotics for AOM before and after the intervention | High | Computerised DSS | Yes |
| Coenen (2004) | Belgium | Adults with acute cough | Educational outreach visit (academic detailing) and postal reminder | All intervention and controls received a public campaign | Total antibiotic use, measured as antibiotic prescribing rate by GP for adults with acute cough | Unclear | Educational Outreach | Yes |
| Cohen (2000) | France | Children ≤10 years old with nasopharyngitis | Printed educational material about the management of paediatric nasopharyngitis following a national consensus. Participating clinicians were also encouraged to share this information with parents | Usual care | Total antibiotic use, measured as rate of patients prescribed | High | Printed Educational Material | Yes |
| Cohen (2007) | France | Children with influenza | Rapid test for influenza detection | Usual care | Rate of antibiotic prescribing measured as percentage of patients prescribed, both antibiotics and antivirals | High | POCT Antigen Detection | No |
| Curtis (2021) | UK | NHS General Practices | 3 arm trial (2 active arms):<br>- Feedback (plain) monthly for 3 months by letter, fax and email<br>- Feedback and behavioural impact optimization for engagement | Usual Care | Percentage difference in broad-spectrum antibiotic prescribing during the follow-up periods (5 weeks post each wave of intervention n=3) | High | Audit and Feedback | No |
| DahlerEriksen (1999) | Denmark | All patients | Point of Care Testing C-Reactive Protein | Crossover trial | Total antibiotic use measured as the rate of patients prescribed in each group | High | POCT Inflammatory Markers | Yes |
| De Santis (1994) | Australia | Patients with tonsillitis | Pharmacist-led educational outreach visit to GPs to discuss tonsillitis management | Not specified | Specific antibiotic use, measured as the percentage of antibiotic prescriptions compliant with guidelines | High | Educational Outreach | No |
| Diederichsen (2000) | Denmark | All ages, consulting for RTI | Point of Care Testing C-Reactive Protein | Usual care | Total antibiotic use measured as the rate of patients prescribed in each group | High | POCT Inflammatory Markers | Yes |
| Dowell (2001) | UK | Adults with respiratory tract infections | Delayed prescribing | Usual care | Collection of antibiotic prescriptions by the patients | Unclear | Other | No |
| EnriquezPuga (2009) | UK | All patients | Two outreach visits | Control practices had educational outreach about antidepressant use | Specific antibiotic use (co-amoxiclav and quinolones), measured as both:<br>- Number of prescriptions per 1000 registered patients.<br>- % co-amoxiclav and quinolones, of total antibiotic prescribing | Unclear | Educational Outreach | No |
| Figueiras (2020) | Spain | Primary care physicians | One-hour educational outreach visits with optional use of computerised DSS | Usual care | Outpatient antibiotic use presented as defined daily doses per 1000 inhabitants per day (DID) | High | Educational Outreach | Yes |
| Filkenstein (2001) | U.S. | Children <6 years old | Two educational meetings with a clinician peer leader using CDC summaries about prudent AB prescribing | Usual care | Total antibiotic use, measured as antibiotic courses per person-year | Low | Educational Outreach | Yes |
| Fleet (2014) | UK | Nursing home residents | RAMP antibiotic stewardship tool (printed): checklist to complete | Usual care | Total antibiotic use measured as number of prescriptions per 100 residents | High | Printed DSS | Yes |
| Flottorp (2002) | Norway | Sore throat in people aged 3 years or older.<br>Urinary tract infection in non-pregnant women aged 16 to 55 | Multicomponent:<br>- DSS<br>- Education for GPs and patients<br>- Printed material<br>- Financial incentive | Usual care for either sore throat or UTI (each active arm is the control for the condition they are not targeted for) | Total antibiotic use, measured as the rate of patients prescribed antibiotics | Unclear | Multifaceted<br>- Other | Yes |
| Forrest (2013) | U.S. | Children aged 2 months to 12 years with acute otitis media (AOM) | 2x2 factorial trial:<br>- DSS<br>- Audit and feedback<br>- DSS plus audit and feedback<br>- Usual care | Usual care | Percentage of visits in which amoxicillin (first line) was prescribed | High | Multifaceted<br>- Other | No |
| Francis (2009) | UK | Children with respiratory tract infections | Interactive booklet | Usual care | Total antibiotic use, measured as rate of patients taken antibiotics at two weeks | Unclear | Other | No |
| Gerber (2013) | U.S. | Children aged 1 to 10 years old with acute respiratory tract infection | One-hour educational session plus feedback about current prescribing at practice and individual prescriber level | Usual care | Proportion of broad-spectrum antibiotic prescriptions | Unclear | Multifaceted<br>– Education with Audit and Feedback | Yes |
| Gjelstad (2013) | Norway | Patients with acute respiratory tract infections | Two academic detailing visits, reviewing antibiotic guidelines and each GP's prescribing | Control practices received education about inappropriate prescribing in the elderly | Total antibiotic prescribing rates, expressed as number of antibiotic prescriptions for number of episodes of respiratory tract infections | Low | Educational Outreach | No |
| Gonzales (2013) | U.S. | 13 years old and older with acute uncomplicated bronchitis | 3 arm trial (2 active arms):<br>- Printed DSS<br>- Computerised DSS | Usual care | Total antibiotic prescribing rates for acute bronchitis measured as the percentage of patients with acute bronchitis prescribed antibiotics | Low | Printed DSS<br>Computerised DSS | Yes |
| Hurlimann (2015) | Switzerland | Patients with respiratory tract infections | Audit and feedback on with guidelines sent to all prescribers | Usual care | Rate of patients prescribed specific antibiotic classes | Low | Audit and Feedback | No |
| Hux (1999) | Canada | Patients over 65 years old with respiratory and urinary tract infections | Posted feedback was provided to prescribers three times in a six months' period about their own individual prescribing practices and how they compared to a group of peers, along with guidelines | Usual care | Proportion of visits in which specific antibiotics were used | Unclear | Audit and Feedback | No |
| Ilet (2000) | Australia | Patients with respiratory or urinary tract infections | 10-15 minutes visit by adviser to individual prescribers to discuss recommended treatments for respiratory tract infections | Usual care | Total antibiotic use, measured as median number of antibiotic prescriptions by GP | High | Educational Outreach | No |
| Keitel (2017) | Tanzania | Patients aged between 2 and 59 months being treated with febrile illnesses | Multifaceted - Decision Support System (DSS) Computerised based on C-Reactive Protein and procalcitonin point of care testing | Electronic algorithm derived from Integrated Management of Childhood Illness (IMCI) strategy (developed by, WHO and UNICEF). | The proportion of antibiotic prescriptions at day 0. Prescribing between day 1 and day 6 were recorded but involved non-study physicians | High | Multifaceted other | Yes |
| Lagerlov (2000) | Norway | People with urinary tract infection | 2 educational evening meetings, concerning diagnosis and management | Usual care | Rates of “acceptable” and “unacceptable” antibiotic prescribing. The difference in proportions of short (acceptable) and long (unacceptable) treatments for UTI before and after the intervention | Low | Educational Workshop | No |
| Le Corvosier (2013) | France | People with respiratory tract infection | Interactive seminar presenting evidence-based guidelines on antibiotic prescription for respiratory infections | Usual care | Total antibiotic use, measured as the proportion of prescriptions (of total) that were antibiotics (meta-analysed) | Unclear | Educational Workshop | Yes |
| Linder (2009) | U.S. | Patients with acute respiratory tract infection | Electronic health record integrated decision support tool providing guidance and treatment options for respiratory tract infections - ‘ARI Smart Form’ | Usual care | Antibiotic prescribing rate expressed as percentage of patients consulting with acute respiratory tract infections who are prescribed antibiotics | High | Computerised DSS | Yes |
| Little (2013a) | UK | Patients aged three years old or older with acute sore throat | Clinical score plus CRP POCT | Usual care | Rate of patients prescribed antibiotics | Low | POCT Inflammatory Markers<br>Multifaceted - Other | Yes |
| Little (2013b) | UK | Patients with respiratory tract infection | 2x2 factorial trial:<br>- POCT - CRP<br>- Education<br>- POCT - CRP plus education | Usual care | Total antibiotic use measured as the rate of patients prescribed in each group | Unclear | Educational Workshop | Yes |
|  |  |  | - Usual care |  |  |  |  |  |
| Little P (2014) | UK | Patients aged three years old and over with respiratory infection | Delayed prescribing | Immediate antibiotic use | Rate of patients using antibiotics | Unclear | Other | No |
| Llor (2011) | Spain | Patients aged 14 to 60 years old with acute pharyngitis | Rapid test for Group A *Streptococcus* | Usual care | Antibiotic prescription rate measured as the rate patients with pharyngitis prescribed antibiotics | Unclear | POCT Antigen Detection | Yes |
| Loeb (2005) | Canada and U.S. | Nursing home residents with urinary tract infection | 30-minute visit to both nurses and GPs presenting case scenarios and a clinical algorithm | Usual care | Total antibiotic use, measured as number of antibiotic courses per 1000 residents-day | Unclear | Educational Outreach | Yes |
| Lundborg (1999) | Sweden | Women aged 18 to 75 years old with urinary tract infection | Outreach visits on urinary tract infections, using feedback as learning tool in peer groups | Control practices received education on asthma | Total antibiotic use, measured as number of prescriptions for UTI (as % of total antibiotic use) | Unclear | Educational Outreach | Yes |
| Martens (2007) | The Netherlands | All patients | Computerised prescription model as reminder system tool with integrated prescribing guidelines | Usual care | Total antibiotic use measured as the percentage of visits with antibiotic prescribed) | High | Computerised DSS | Yes |
| McConnell (1982) | U.S. | Patients with upper respiratory tract infection | 30 minute educational visits by expert clinicians | Not specified | Specific antibiotic use, measured as mean number of prescriptions of tetracyclines | High | Educational Outreach | No |
| Meeker (2014) | U.S. | Adult (over 18 years old) patients with | Display of poster-sized 'commitment letters' in doctors' offices | Usual care | Rate (%) of patients prescribed 'inappropriate' antibiotics for respiratory tract infections | Low | Other | No |
| Milos (2013) | Sweden | Patients with respiratory tract infections, all ages and aged 0-6 years | Three arm trial comparing two behavioural interventions | Usual care | Total antibiotic use measured as number of antibiotic prescriptions for RTIs per 1000 registered patients | Unclear | Other | No |
| Naughton (2009) | Ireland | All patients | Half hour educational outreach visit versus postal feedback | Postal feedback | Total antibiotic prescribing measured as prescriptions per 1000 registered patients | Unclear | Other | No |
| O'Connell (1999) | Australia | All patients | Individualised text and graphical feedback to prescribers about their prescribing rates of five main drug groups | Usual care | Total antibiotic prescribing measured as number of prescriptions per 100 services | Low | Audit and Feedback | No |
| Pagaiya (2005) | Thailand | All patients | Educational out-reach visit about antibiotic use in respiratory tract infections and diarrhoea | Usual care | Total antibiotic use, expressed as percentage of patients prescribed antibiotics | High | Educational Outreach | Yes |
| Rebnord (2016) | Norway | Participants aged between 0-6 years old | Point of care testing C-Reactive Protein | Usual care | Total antibiotic use measured as the rate of patients prescribed in each group | High | POCT Inflammatory Markers | Yes |
| Regev-Yochay (2011) | Israel | Children (younger than 18 years old) | Multifaceted intervention over 3 years. Comparisons included:<br>- Educational workshop (end of year 2) versus usual care<br>- Audit and feedback (year 3 compared to year 2)<br>- Mixed audit and feedback plus education (year 3 compared to baseline) | Usual care | Total antibiotic use, measured as the number of prescriptions per 100 patient-years | Low | Educational Workshop<br>Audit and Feedback<br>Multifaceted – Education with Audit and Feedback | Yes |
| Samore (2005) | U.S. | Patients with acute respiratory tract infection | DSS (printed and computerised) plus professional education in practices plus a community intervention to change help seeking behaviour (one comparison from three arm study where the second active arm only received the community intervention) | Usual care | Total antibiotic use measured as the number of prescriptions per 100 patients-years | Unclear | Multifaceted - Other | Yes |
| Santoso (1996) | Indonesia | Patients with diarrhoea | 3 arm trial (2 active arms):<br>- Educational seminar<br>- Small group face-to-face educational outreach | Usual care | Total antibiotic use, measured as percentage of patients prescribed antibiotics (meta-analysed) | High | Educational Workshop<br>Educational Outreach | Yes |
| Shrestha (2006) | Nepal | Adults with respiratory tract infection | Educational outreach visits to health workers presenting WHO guidelines of “Practical Approach to Lung Health” (PAL) | Usual care | Total antibiotic use, measured as % encounters in which antibiotic is prescribed | High | Educational Outreach | No |
| Sondergaard (2003) | Denmark | Any age with respiratory tract infections | Dissemination of written clinical guidelines with feedback | Usual care | Total antibiotic prescription rate, measured as mean number of antibiotic prescriptions per 1000 registered patients | Unclear | Audit and Feedback | Yes |
| Van Den Bruel (2016) | UK | Participants aged between 1 month and 16 years | Point of care testing C-Reactive Protein | Usual care | Total antibiotic use measured as the rate of patients prescribed in each group | High | POCT Inflammatory Markers | Yes |
| Van Driel (2007) | Belgium | Patients with rhinosinusitis | Use of quality circles to promote and discuss guidelines | Usual care | Total antibiotic use measured as rate of patients receiving antibiotics | High | Other | No |
| Varonen (2007) | Finland | Patients with sinusitis | Implementation of national guidelines with practices randomised to one of two active arms<br>- academic detailing<br>- problem-based learning | Two active arms compared | Specific antibiotic use. Use of first-line drugs, measured as percentage (%) of courses complying with recommended duration | Unclear | Educational Workshop | No |
| Vellinga (2016) | Ireland | All patients attending one of the 32 eligible Irish Primary Care Research Network practices | Information on national guidelines for antibiotic prescribing with a reminder pop-up of these guidelines. Discussion of the first practice audit report and a monthly audit post intervention | Baseline data collection with a coding workshop to all practices including the control arm | Rate of total antibiotic prescribing measured as percentage of patients prescribed | Unclear | Multifaceted - Education with Audit and Feedback | Yes |
| Veninga (2000) | The Netherlands | Women aged 18 to 75 years old with urinary tract infection | Two educational meetings about guidelines for management of urinary tract infections (UTI) | People with asthma (no UTI intervention) | Specific antibiotic use, measured as the proportion of DDDs which were for first line drugs | Unclear | Educational Workshop | No |
| Wei (2017) | China | Participants aged to 14 years old | Educational material, printed and interactive session | Usual care | Rate of total antibiotic prescribing measured as percentage of patients prescribed | Unclear | Educational material | Yes |
| Welschen (2004) | The Netherlands | Patients of any age with acute respiratory tract infection | Education about first-line antibiotics for respiratory tract infections and enhanced communication, with feedback on prescribing rates. | Usual care | Total antibiotic use, measured as the proportion of practice encounters for respiratory tract infection in which antibiotics are prescribed | Unclear | Multifaceted – Education with Audit and Feedback | Yes |
| Wilson (2003) | Australia | Children with respiratory tract infections | Intensive intervention with focus groups and workshops for guideline development in the first year, and educational outreach with prescribing feedback and reinforcement of guidelines in the second year | Control practices also received the guidelines and an implementation package | Total antibiotic use, measured as the number of subsidised antibiotic prescriptions per 100 Medicaid claims | High | Multifaceted -Other | Yes |
| Worrall (2007) | Canada | Adults with acute sore throat | 2x2 factorial trial:<br>- Printed DSS<br>- Rapid test for Group A *Streptococcus* | Usual care | Antibiotic prescribing rate expressed as % visits where antibiotic was prescribed | Unclear | POCT Antigen Detection<br>Printed DSS | Yes |
| Yang (2014) | China | Patients with respiratory tract infections | Public reporting of prescribing rates at both individual and institutional level monthly, with comparison and ranking of institutions, and data available to both patients and providers |  | Proportion of patients (%) with respiratory tract infections receiving antibiotics | High | Other | No |
| Zahlanie (2021) | USA | Patients between the ages of 3 to 19 months who had received prescriptions for oral antibiotics and/or treatment in 4 outpatient clinics in the USA | A quality improvement project evaluating the impact of EPIC order sets (providing options of antibiotics with dosage and duration time) and educational outreach sessions on antibiotic prescribing for common paediatric bacterial ARIs | Usual Care | Percentage of first-line antibiotic prescribing in both arms, pre and post implementation of the intervention | High | Educational Outreach | Yes |
| Zwar (1995) | Australia | URTI and people with tonsillitis | Intervention GP trainees attended educational seminar about antibiotic prescribing | GP trainees who attended a seminar about cancer prevention | Total antibiotic prescribing for any condition and for uncomplicated URIs, measured as the GP trainee antibiotic prescribing rate per patient encounter | High | Educational Workshop | Yes |

### Risk of bias

Overall, 30/74 (41%) studies had high risk of bias, 33/74 (46%) had unclear, and 11/74 (15%) had low (Figure 2, Supplementary Figure S1). The most common sources of high risk of bias were lack of reporting of baseline outcome measures 11/74 (15%), lack of reporting of baseline characteristics 8/74 (11%), lack of blinding, and other bias, both 7/74 (9%). Most studies had low risk of bias in relation to selective outcome reporting 72/74 (97%), completeness of outcome data 72/74 (97%) and allocation concealment 67/74 (91%).

**Figure 2.**
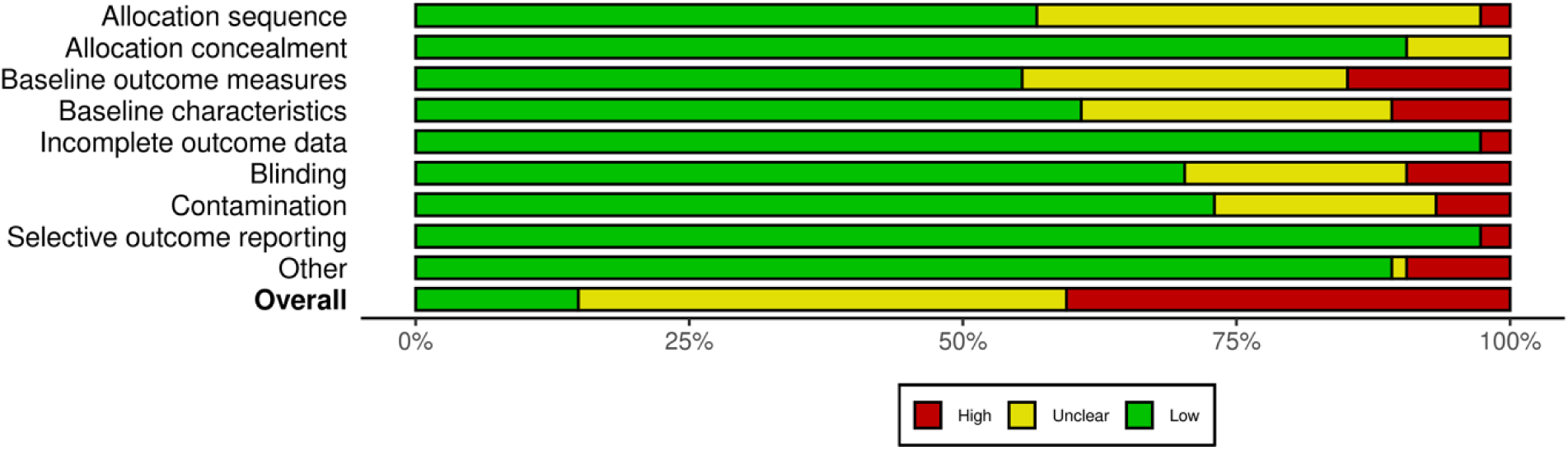
- Overall Risk of Bias – percentage of studies assigned each category

### Intervention effects

Pooled estimates of intervention effects on total antibiotic prescribing across all intervention types are summarised in Table 2 for primary meta-analyses (Figure 3) and sensitivity meta-analyses (Supplementary Figure S2).

**Figure 3.**
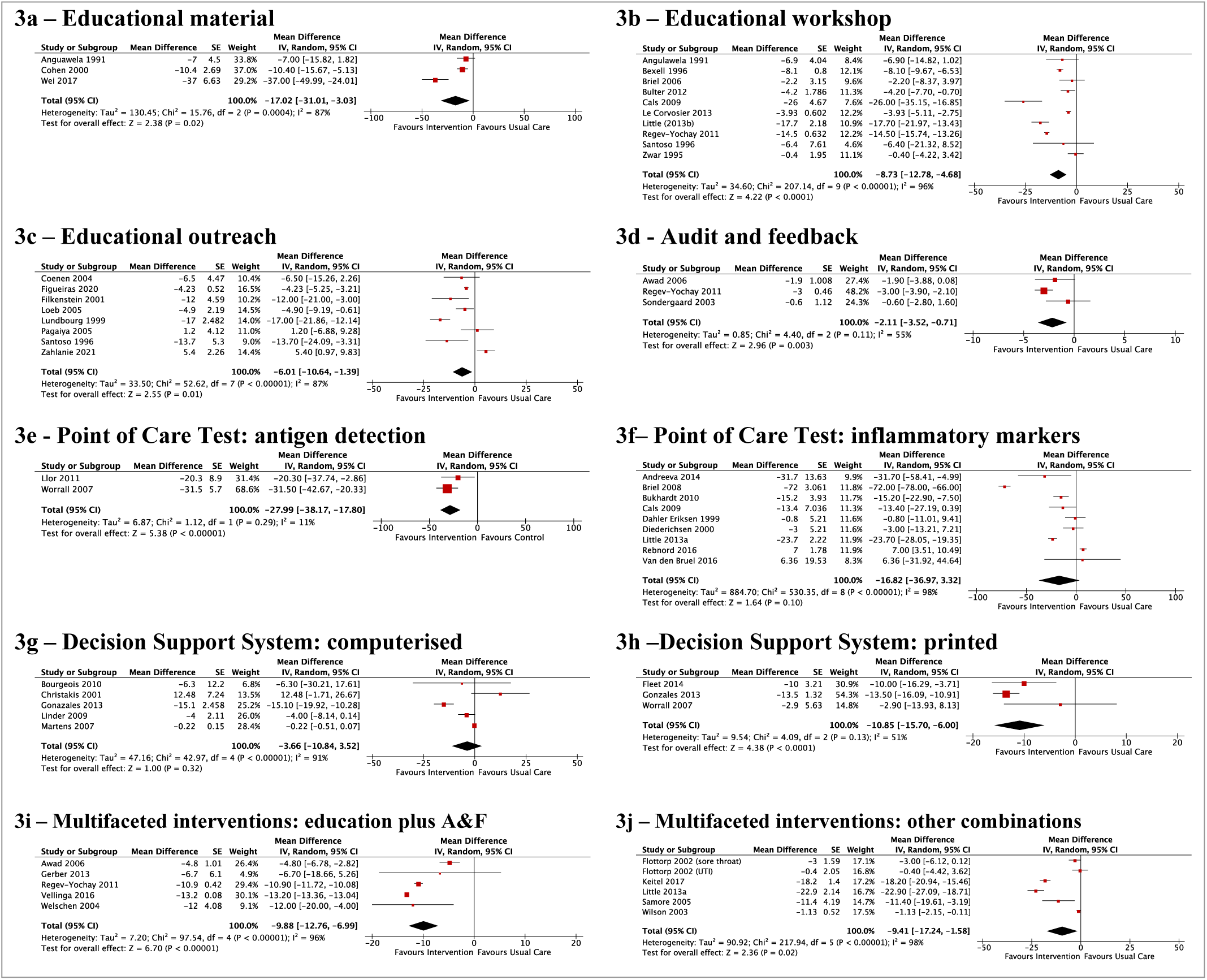
- Forest plots of main intervention effects

**Table 2.**
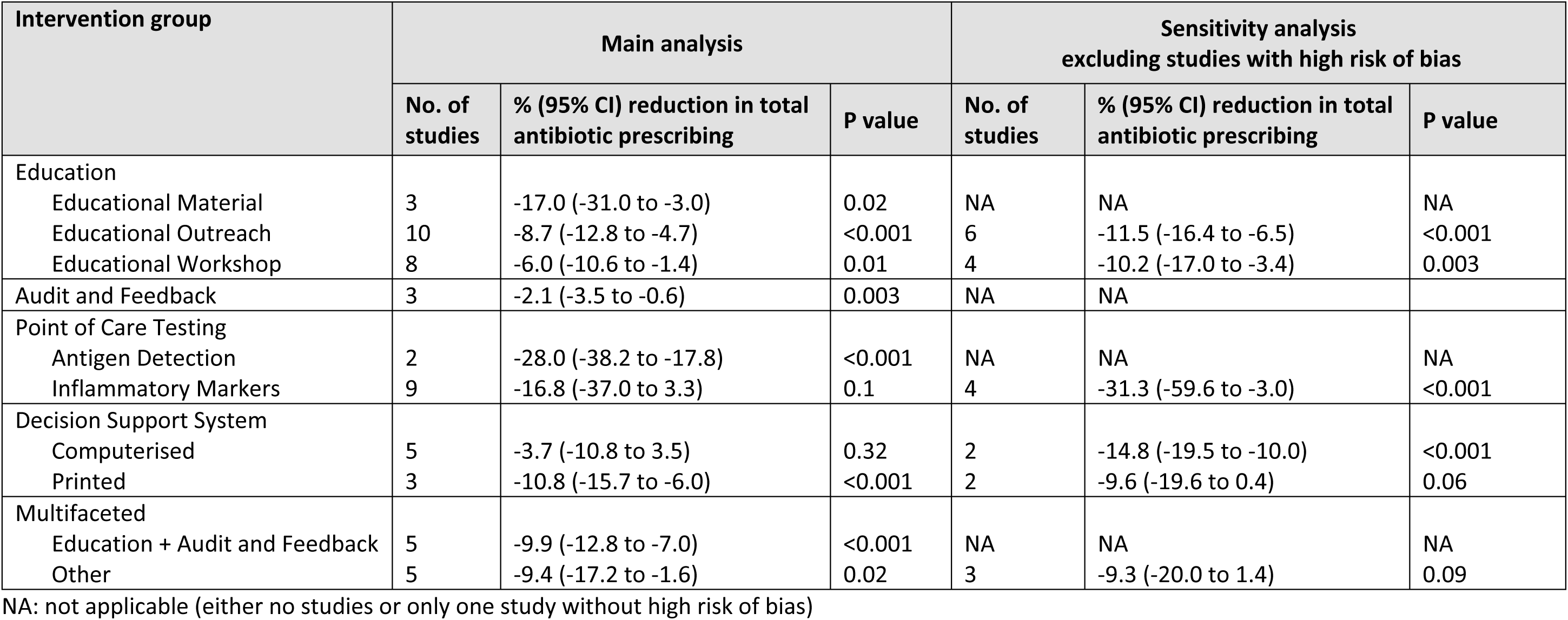
- Pooled meta-analysis estimates of intervention effects on total antibiotic prescribing

#### Education

##### a. Educational Material

Four studies compared educational material with usual care. Two had high risk of bias^28, 29^ and two unclear.^30, 31^ Meta-analysis including three studies^28, 29, 31^ found a large, significant reduction in total antibiotic prescribing (mean percentage difference −17.0% [95%CI −31.0, - 3.0], p=0.02) (Figure 3a). The fourth trial (a three-armed trial with an educational intervention) found no change in targeted prescribing of a single antibiotic.^30^

##### b. Educational Workshops

Fourteen studies compared educational workshops with usual care. Five had high risk of bias,^28, 32–35^ six unclear,^36–41^ and three low.^42–44^ Meta-analysis including ten studies^28, 33–38, 41, 42, 44^ found a moderate, significant reduction in antibiotic prescribing (−8.7% [−12.8, -4.7], p<0.001) (Figure 3b). Sensitivity analysis including six studies^36–38, 41, 42, 44^ found a slightly larger significant reduction (−11.5% [−16.4, -6.5], p<0.001) (Supplementary Figure S2a).

Of the four studies not meta-analysed, two reported significant reductions in antibiotic prescribing for urinary tract infections,^32, 40^ one reported a 13% improvement in the duration of prescribed antibiotic courses,^43^ and one reported increased prescribing of a recommended specific antibiotic.^39^

##### c. Educational Outreach

Sixteen studies compared educational outreach with usual care. Eight had high risk of bias,^34, 45–51^ five unclear,^30, 52–55^ and three low.^56–58^ Meta-analysis including eight studies ^34, 46, 49, 51, 52, 54, 55, 57^ found a small, significant reduction in antibiotic prescribing (−6.0% [−10.6, - 1.4], p=0.01) (Figure 3c). The reduction was larger in sensitivity analysis including four studies^52, 54, 55, 57^ (−10.2 % [−17.0, −3.4], p=0.003) (Supplementary Figure S2b).

Eight studies were not meta-analysed.^30, 45, 47, 48, 50, 53, 56, 58^ Four found significantly lower antibiotic prescribing, with two targeting specific antibiotics^30, 48^ and two targeting respiratory tract infections.^56, 58^ Two studies found an increase in prescribing of recommended specific antibiotics.^45, 47^ Two studies found non-significant reductions in prescribing, one of specific antibiotics^53^ and the other for respiratory tract infections.^50^

#### Audit and Feedback

Seven studies compared audit and feedback with usual care. One had high risk of bias,^59^ three unclear,^60–62^ and three low.^44, 63, 64^ Meta-analysis of three studies ^44, 60, 62^ found a small, significant reduction in antibiotic prescribing (−2.1% [-3.5, -0.6], p=0.003) (Figure 3d).

Four studies were not meta-analysed. Three reported improvements in prescribing rates for specific antibiotics.^59, 61, 63^ One reported no difference in median prescribing rates per 100 consultations.^64^

#### Point of Care Testing (POCT)

##### a. Antigen Detection

Three studies compared POCT for antigen detection with usual care. One had high risk of bias ^65^ and two had unclear risk of bias.^66, 67^ Meta-analysis of two studies ^66, 67^ found a large, significant reduction in antibiotic prescribing (–28.0% [-38.2, −17.8], p<0.001) (Figure 3e).

In the study not meta-analysed,^65^ use of POCT for flu in children increased antibiotic use (9.5% of patients with test performed *versus* 3.9% in those without), particularly among those with a negative test (15.7% *versus* 4.3% of those with a positive test).

##### b. Inflammatory Markers

Nine studies compared POCT for inflammatory markers with usual care. Five had high risk of bias,^68–72^ three unclear^37, 73, 74^ and one low.^75^ Meta-analysis including all nine studies^37, 68–75^ found no significant reduction in antibiotic prescribing (−16.8% [-37.0, 3.3], p=0.10) (Figure 3f), but in sensitivity analysis of four studies^37, 73–75^ the reduction was larger and statistically significant (−31.3% [-59.6, −3.0], p<0.001) (Supplementary Figure S2c).

#### Decision Support Systems (DSS)

##### a. Computerised

Five studies compared computerised DSS with usual care. Three had high risk of bias,^76–78^ one unclear,^79^ and one low.^80^ Meta-analysis of all five^76–80^ found a small non-significant reduction in prescribing (−3.7% [−10.8, 3.5], p=0.32) (Figure 3g). Sensitivity analysis of two studies^79, 80^ found a large, significant reduction (−14.8% [−19.5, −10.0], p<0.001) (Supplementary Figure S2d).

##### b. Printed

Three studies compared printed DSS with usual care. One had high risk of bias,^81^ one unclear.^67^ and one low.^80^ Meta-analysis of all three studies^67, 80, 81^ found a significant reduction (−10.8% [−15.7, -6.0], p<0.0001) (Figure 3h). Sensitivity analysis including two studies^67, 80^ found a non-significant reduction of similar magnitude (−9.6% [−19.6, 0.4], p=0.06) (see Supplementary Figure S2e).

#### Multifaceted Interventions

##### a. Education with Audit and Feedback

Five studies compared education and audit and feedback with usual care. Four had unclear risk of bias,^60, 82–84^ and one low.^44^ Meta-analysis of all five studies^44, 60, 82–84^ identified a significant reduction in total antibiotic prescribing (−9.9% [−12.8, -7.0], p<0.001) (Figure 3i).

##### b. Multifaceted Other

Six studies reporting seven interventions compared multifaceted interventions other than education with audit and feedback, with usual care. The interventions included various combinations of education for prescribers and patients, audit and feedback, DSS, POCT, and financial incentives (Table 1). Three had high risk of bias^85–87^ and three had unclear.^75, 88, 89^. Meta-analysis of five studies^75, 86–89^ identified a significant reduction (−9.4% [−17.2, −1.6], p=0.02) (Figure 3j). In sensitivity analysis including three studies,^75, 88, 89^ the reduction was the same magnitude but no longer significant (−9.3% [-20.0, 1.4], p=0.09) (Supplementary Figure S2f). The study that was not meta-analysed found a combination of audit and feedback and computerised DSS did not change amoxicillin prescribing rates.^85^

#### Other Interventions

Twelve included studies did not fit into the above intervention groups. Three had high risk of bias,^90–92^ eight unclear ^93–100^ and one low.^101^

One study^94^ reported longer term follow-up (avg. 3.7 years later) of another included study.^37^ The follow up data could not be extracted or included seperately, but the previously reported significant reduction in antibiotic prescribing was not sustained.

Three studies examined delayed prescribing, two compared this with usual care for respiratory tract infections and found significant reductions in antibiotic use,93, 96 and one compared it with decision support and found no significant difference.98

One study compared different types of education and reported a significant reduction in total antibiotic use,95 while another compared educational outreach with postal feedback and reported no difference.100 Two studies examined novel ways of guideline dissemination, using text messages90 and quality circles,91 but neither had significant effects.

Three studies reported interventions using behavioural techniques, with two finding significant effects on the targeted prescribing for respiratory tract infections,97, 101 and one finding significant effects in paediatric patients <6 years only but no effect on the general population.^99^ Finally, one study examined public reporting of primary care antibiotic prescribing and found a significant effect.^92^

## Discussion

### Principal Findings

This systematic review with meta-analyses of randomised controlled trials in community settings found statistically significant reductions in total antibiotic prescribing with the following interventions compared to usual care: educational material (−17.0%); educational workshops (−8.7%); educational outreach (−6.0%); audit and feedback (−2.1%); POCT for antigen detection (−38.2%), and; printed DSS (−10.9%). Multifaceted interventions incorporating education with audit and feedback (−9.9%) and other combinations of interventions (−9.4%) also significantly reduced total antibiotic prescribing by similar magnitudes as single interventions (Figure 3, Table 2). Sensitivity analyses, excluding studies with high risk of bias, had similar findings except that the effects of POCT for inflammatory markers (−31.3%) and computerised DSS (−14.8%) were statistically significant, and printed DSS and multifaceted ‘other’ were no longer statistically significant (Figure S2, Table 2).

### Strengths and Limitations

Strengths of the study include the use of a comprehensive search carried out in MEDLINE, EMBASE and The Cochrane Effective Practice and Organisation of Care (EPOC) Specialised Register, aiming to identify all potentially relevant studies. No language restrictions were placed and included studies included low- and middle-income countries. The mean percentage difference in total antibiotic prescribing and the generic inverse variance method was used for meta-analysis because of variability in outcome reporting. In this method, larger studies with smaller standard errors are given greater weight, improving the precision of the effect estimate.

A limitation is the heterogeneity among studies, in trial design, interventions, and outcome measures. Analysis therefore grouped similar intervention types and used random-effects meta-analysis. There was still heterogeneity within groups and some studies being ‘mixed’. Classification of some studies by intervention type required careful consideration and discussion but a dominant intervention could usually be identified for study allocation.

There will be variation in context and implementation across studies but this is not possible to account for in meta-analyses and is a common limitation across any evidence synthesis involving behavioural type interventions.

### Comparison with other studies

All studies from the 2005 Arnold *et al* Cochrane review of primary care stewardship interventions were assessed for eligibility. Of the 40 studies they included, 17 are included and 23 were ineligible, and we included four studies they excluded. Their review included interventions involving printed educational materials, educational meetings, educational outreach visits, audit and feedback, financial and healthcare system changes, physician reminders, patient-based interventions, and multifaceted interventions, with multifaceted interventions the most effective. In our review, multifaceted approaches (categorised as ‘education plus audit and feedback’ and ‘other multifaceted’) were not clearly more effective than single intervention strategies.

The largest effect observed in our review, a -28% reduction in antibiotic use, was with POCT for antigen detection. This intervention was not included in the previous Cochrane review^16^ and we only identified two studies - both targeting patients with suspected respiratory infections. POCT for inflammatory markers had more included studies. We found a non-significant decrease (−17%) in antibiotic prescribing across all nine, but the effect was significant in sensitivity analysis including four high quality trials (−31%) (Table 2). Four recent systematic reviews examined POCT for inflammatory markers in respiratory tract infections. One included different intervention types in primary care,^102^ but two that included POCT tests for any inflammatory markers^22, 23^ each identified 13 studies, with eight overlapping and six of the total combined 18 studies included in our review. Both reported reductions in antibiotic treatment at index consultation with CRP POCT (RR 0.77 [0.69 to 0.86]^22^ and RR 0.79 [0.70 to 0.90]^23^), of similar magnitude to our findings. One also reported a reduction (RR 0.81 [0.76 to 0.86])^22^ at 28 days, but the other reported no difference, and an increase in re-consultation.^23^ The fourth review, of procalcitonin, found only two of 14 eligible RCTs were in primary care.^103^ In those two studies (both in our review),^33, 74^ procalcitonin POCT lowered antibiotic use (OR 0.10 [0.07 to 0.14]). Overall, POCT in primary care stewardship shows promise in trial settings but there has been little (or no) evidence of translation into routine clinical practice.

Educational interventions, most often involving guideline implementation, have been associated with improvements in use and appropriateness of antibiotics in primary care,^20^ but we found an unexpectedly large effect of printed educational materials (−17% reduction), particularly since workshops (−8.7%) and outreach (−6%) had smaller effects. This large effect contrasts with a recent Cochrane review assessing the effect of printed educational materials on professional practice more generally, which reported that they “probably” improve practice compared to no intervention (median absolute risk difference 0.04 [IQR 0.01 to 0.09]).^104^ The differences may be contextual and/or reflect that our meta-analysis included only three studies (compared to 16),^104^ one of which had a very large effect (−37%),^31^ and two had high risk of bias.^28, 29^

The small but significant effects of audit and feedback on total antibiotic prescribing in our review (−2% reduction) are consistent with a broader systematic review of the effects of audit and feedback on healthcare professional practice reporting a risk difference of 4.3% (IQR 0.5 to 16) across 49 studies with dichotomous outcomes and percent change of 1.3% (IQR = 1.3% to 28.9%) across 21 studies with continuous outcomes.^105^

A systematic review of computerised decision support systems (DSS) to reduce inappropriate antibiotic prescribing included non-randomised studies and hospital settings, in addition to RCTs in primary care settings.^106^ Of 13 RCTs (four included in our review^36, 76, 77, 80^) only two, in hospitals, were included in their meta-analysis. Similar to our review, they report a non-significant effect favouring DSS (OR 1.24 [0.95 to 1.62]).^106^ We did, however, find, a significant reduction in antibiotic prescribing (−14.8%, p<0.001) when studies with high risk of bias were excluded (Table 2).

A Cochrane review of 221 studies of antibiotic stewardship in hospital settings included non-randomised studies and clinical and unintended outcomes in addition to prescribing outcomes, and grouped studies for meta-analysis by outcome with meta-regression to estimate the effect of behaviour change functions.^19^ It reported a 15% (14 to 16) change in the intended direction across 29 RCTs with a prescribing outcome, and that enablement (defined as ‘increasing means/reducing barriers to increase capability or opportunity’), restriction (‘using rules to reduce the opportunity to engage in the target behaviour [or increase the target behaviour by reducing the opportunity to engage in competing behaviours]’), and targeting antibiotic choice over exposure were significantly associated with greater intervention effect.^19^ Although not directly comparable to our primary care review, the overall conclusion that most published stewardship interventions are effective, with some variation in effect size by type of intervention, is consistent with our findings.

### Implications for clinicians and policy makers

The evidence synthesised in this review indicates that a wide range of stewardship interventions are effective in reducing antibiotic use in primary care, but defining which intervention component is most effective is complex. With increasingly challenging targets for reductions in antibiotic prescribing in primary care,^107^ and AMR a high priority,^108^ there is a need for continued implementation of stewardship interventions. Sustained stewardship programmes in primary care, with sequential interventions and a likely gradual change in prescribing culture, have been effective in reducing total antibiotic prescribing at national level over time,^109–112^ and more focused interventions can have large effects on the choice of antibiotic prescribed in primary care over shorter time periods.^113^ Designers of antimicrobial stewardship programmes can choose from a range of likely effective intervention components that are appropriate to their context in terms of local feasibility and acceptability.

### Implications for future research

Further RCTs of single stewardship interventions are unlikely to significantly add to the evidence, given that, overall, all types of intervention were effective in improving antibiotic prescribing. Better designed comparisons between different multifaceted interventions are likely to be more impactful, given the large effects of multifaceted interventions in real-world analyses,^113^ although we did not observe this effect in the small number of multifaceted trials in this review. Improving study quality and standardising outcome measures would facilitate more informative evidence synthesis to compare interventions.

Pragmatic trials of more real-world implementation are also needed to evaluate interventions such as POCT that are promising in trials but have not been adopted into practice. POCT had the largest effect size in this review, but the meta-analyses included small numbers of studies and practices participating are unlikely to be representative of the wider prescriber population. Previously reported barriers to implementation of POCT in primary care included the impact on staff and workflow, as well as concerns around cost, reliability/quality control and patient perception.^114^ The widespread use of lateral flow testing in the COVID−19 pandemic may have had an influence on acceptability but concerns about the impact on primary care workflow, with appointments typically less than 10 minutes and an increase in telephone consultations in the UK, are likely to persist. Implementation research in this area should be high priority.

More research is also needed on the effect of stewardship interventions on outcomes other than prescribing.^115^ Outcomes should include intended outcomes such as reductions in AMR^7^ and *Clostridioides difficile* infection,^116^ and unintended consequences of reduced prescribing such as hospital admissions.^117^ These can all be more challenging to evaluate in relatively small RCT populations, with small numbers of outcomes, than in evaluations of real-world interventions across whole populations. Non-randomised evaluation of the effect of real-world stewardship interventions, including multi-faceted stewardship interventions, on prescribing and other outcomes should be prioritised as they can generate valuable evidence.^7, 115–117^ However, rigorous evaluation requires more systematic documentation of the components included in real world stewardship interventions.

## Conclusions

Published trials of all types of stewardship interventions in primary care were effective in improving antibiotic prescribing. More pragmatic evaluations of multifaceted interventions and technologies in ‘real-world’ practice could inform the design of future stewardship programmes.

## Contributorship Statement

Virginia Hernandez Santiago (VHS) and Rebecca Andrews (RA) contributed equally to this paper and work.

VHS, BG and CM designed the work. All authors contributed to selection and review of studies for inclusion, and risk of bias assessment. RA and VHS carried out data extraction and meta-analysis, with quality checking and resolution of uncertainty by BG and CM. VHS and RA wrote the first draft of the manuscript with all authors contributing to critical review and writing the final version. The corresponding author attests that all listed authors meet authorship criteria and that no others meeting the criteria have been omitted.

## Ethics approval

Not required.

## Transparency statement

VHS, as the guarantor, affirms that the manuscript is an honest, accurate, and transparent account of the work reported; that no important aspects have been omitted; with no discrepancies from the study as originally planned.

## Data Availability

All data are contained within the manuscript, supporting files and references.

## Supporting information

Supplementary Material file which includes:

- Table S1 - Complete Search Strategy

- Table S2 - Cochrane EPOC group risk of bias criteria for RCTs and QRCTs

- Table S3- Deviations from original PROSPERO registration

- Figure S1 - Risk of Bias in Included Studies

- Figure S2 - Forest plots of sensitivity analyses of main intervention effects

